# Mendelian Randomization Reveals Causal Pathways Between Plasma Protein Quantitative Trait Loci and Graves’Disease Mediated by Immune Cells

**DOI:** 10.1101/2025.01.02.25319932

**Authors:** Shixian Cui, Qingyang Liu

**Affiliations:** The Department of Endocrinology, Liaoning University of Traditional Chinese Medicine Affiliated Hospital,Shenyang,Liaoning,China

**Keywords:** Graves’ disease, Plasma protein quantitative trait loci, Mendelian randomization, Immune cells, Autoimmune disorders, Genetic association

## Abstract

Graves’ disease (GD) is an autoimmune disorder characterized by hyperthyroidism caused by thyroid-stimulating antibodies (TRAb) activating the thyroid-stimulating hormone receptor (TSHR). Current treatments, including antithyroid drugs, radioactive iodine therapy, and surgery, can control hormone secretion but fail to prevent the immune system’s persistent attacks on the thyroid gland. This highlights the need for in-depth research into the immune pathogenesis of GD to develop novel therapeutic strategies.

The study results showed that several key pQTLs, including ACE, PMM2, FCRL4, and DPEP1, are significantly associated with GD. Further investigation revealed that ACE inhibitors (ACEIs) are potentially relevant for GD treatment. These inhibitors are not only widely used in the treatment of hypertension and cardiovascular diseases but also exhibit immune-regulatory effects. For example, by modulating the activity of T cells and B cells, ACEIs have demonstrated anti-inflammatory effects in models of various immune-related diseases.

This study is the first to combine pQTLs with immune cell regulatory mechanisms to explore their impact on the pathogenesis of GD. By integrating genetic variants, immune cell phenotypes, and causal relationship analysis, we not only validated previously reported GD-related pQTLs but also identified new candidate signals and pathways. These findings deepen our understanding of GD pathogenesis and provide a foundation for developing precision therapies and combination treatments targeting GD and related autoimmune diseases.

## Introduction

Graves’ disease is an autoimmune disorder characterized by hyperthyroidism and is one of the leading causes of thyroid dysfunction. The underlying mechanism involves the activation of the thyroid-stimulating hormone receptor (TSHR) by thyroid receptor antibodies (TRAb), resulting in excessive secretion of thyroid hormones, which accelerates metabolism and triggers a range of systemic symptoms. A subset of patients also develops Graves’ orbitopathy(GO), manifested by symptoms such as exophthalmos and visual disturbances [1].

Current treatment options include antithyroid drugs, radioactive iodine therapy, and surgical resection, all aimed at controlling thyroid hormone secretion. However, these treatments do not provide a cure for the disease and are unable to prevent ongoing immune system-mediated attacks on the thyroid. In some cases, the therapeutic response is limited, and patients may require long-term thyroid hormone replacement following radioactive iodine treatment or surgery. Therefore, a deeper understanding of the immune pathogenesis of GD is crucial for the development of novel therapeutic strategies [2].

Recent advancements in genome-wide association studies (GWAS) have identified thousands of plasma protein quantitative trait loci (pQTL) [3]. These studies offer new avenues for exploring the causal relationships between plasma proteins and GD, facilitating the identification of potential biomarkers and the evaluation of risk factors associated with the disease. Immune cells also play a central role in the pathogenesis of GD. B cells produce TRAb, which directly activates TSHR, leading to excessive thyroid hormone production. T cells, particularly helper T cells, release pro-inflammatory cytokines that further exacerbate the immune-inflammatory response in the thyroid [4]. Thus, studying the relationship between pQTL, immune cells, and GD is of significant importance.

Mendelian randomization (MR) is a statistical method that uses genetic variants, such as single nucleotide polymorphisms (SNPs), as instrumental variables(IVs) to assess potential causal relationships between exposures and outcomes. MR analysis minimizes the impact of confounding factors and provides robust evidence for causal inference. Through MR, the causal relationship between pQTL and GD can be explored, as well as the mediating role of immune cells in this process [5].

However, to date, no systematic studies have been conducted to investigate how pQTL may mediate GD through immune cell regulation. Furthermore, the direct effects of pQTL and immune cells on GD remain inadequately explored. In this study, we employ a mediational Mendelian randomization approach to examine the mediating effect of immune cells in the relationship between pQTL and GD, while also conducting a preliminary analysis of the direct impact of pQTL and immune cells on the disease. As an exploratory study, our goal is to identify potential causal pathways and candidate signals, providing a foundation for future validation studies and targeted therapeutic development.

## Methods

### Data sources

The pQTL data were sourced from the Icelandic deCODE Genetics study led by Ferkingstad et al., which included an analysis of 4,907 plasma proteins in a cohort of 35,559 Icelandic individuals and identified over 272 million genetic variants(grch37.ensembl.org/Homo_sapiens/Info/Index)[6].

We extracted IVs for GD from FinnGen (https://r11.finngen.fi/).The study included 453, 733 European participants, consisting of 3,437 cases and 450,296 controls[7].

The dataset used in this study, consisting of 731 immunophenotypes (ID: ebi-a-GCST90001391 to ebi-a-GCST90002121) sourced from the GWAS Catalog (https://www.ebi.ac.uk/gwas/), was obtained from 3,757 Europeans and covers a wide range of immune phenotypes, including absolute and relative cell counts, median fluorescence intensities indicating surface antigen expression, and various morphological features[8].

### Selection of instrumental variables

To investigate the genetic basis of pQTLs, we aimed to establish a strong link between these instrumental variables and the exposure variable, specifically protein abundance in our study. This selection adhered to a rigorous p-value threshold of less than 5 × 10⁻⁸, with a minor allele frequency (MAF) between 1% and 99%. Finally, we carefully selected cis-SNPs located within 1 megabase (Mb) of the gene encoding each target protein, chosen to ensure both statistical relevance and a meaningful biological influence on protein levels.

For the instrumental variables used in both immune phenotypes and GD in the reverse causation analysis, our study applied a genome-wide significance threshold of p < 1 × 10⁻⁵ to identify SNPs with strong associations.

The aggregation approach, set with parameters of r2 =0.001 and a window of 10,000 kb, was applied to remove linkage disequilibrium (LD) among the SNPs. To prevent potential biases, palindromic SNPs were excluded. Lastly, the strength of the selected IVs was assessed by calculating the variance (R2) and F-statistics. R2 = [2 × β2 × EAF × (1 − EAF)]/[2 × β2 × EAF × (1 − EAF) + 2 × SE2 × N × EAF × (1 − EAF)], F = [R2 × (N − 2)]/(1 − R2).In this context, β denotes the effect size of the SNP on the phenotype, N is the sample size, EAF stands for the effect allele frequency, and SE represents the standard error associated with the β estimate. Strong evidence for reducing weak instrument bias was observed, as indicated by an F-value of 10[9]. Detailed information can be found in Tables S1-S3.

### Mendelian randomization with mediation analysis

To establish strong causal inference, IVs must satisfy three key conditions: they should be directly linked to the exposure factors, remain uncorrelated with confounders influencing both the exposure and the outcome, and influence the outcome solely through their impact on the exposure, without alternative pathways[10].

We conducted Mendelian Randomization (MR) analysis using the R package “TwoSampleMR” (version 0.5.7), employing the Inverse Variance Weighted (MR-IVW) method by default. A p-value less than 0.05 was considered statistically significant[11]. The weighted median method was used as a supplementary approach. When heterogeneity was detected, the weighted median method adjusted for the influence of invalid IVs, providing robust estimates even when up to 50% of the IVs were invalid[12]. Thus, even in the presence of heterogeneity, the weighted median method could still offer relatively robust estimates of causal effects.

Considering that the accuracy of the IVW method is based on the assumption of no horizontal pleiotropy, we performed MR-Egger regression throughout the MR process to calculate the intercept and conducted horizontal pleiotropy tests to further evaluate robustness. Only results with a p-value greater than 0.05 were considered to indicate no pleiotropic effects[13]; otherwise, the results were excluded from the analysis. Additionally, it is important to note that if the number of SNPs is fewer than three, horizontal pleiotropy testing cannot be performed. To ensure the robustness of the results, such cases were also excluded from the analysis. Subsequently, heterogeneity between IVs was assessed using the Cochran’s Q test based on both the IVW and MR-Egger methods. Leave-One-Out analysis was also conducted as part of our sensitivity analyses.

The mediation analysis was conducted in three sequential steps. The first step assessed the effect of positive pQTLs on positive immunophenotypes, resulting in the estimation of beta1. The second step evaluated how the immunophenotypes identified in the first step influenced GD, leading to the calculation of beta2. The third step measured the overall effect of pQTLs on GD, recorded as betaAll (as shown in Fig 1).

**Fig 1.**
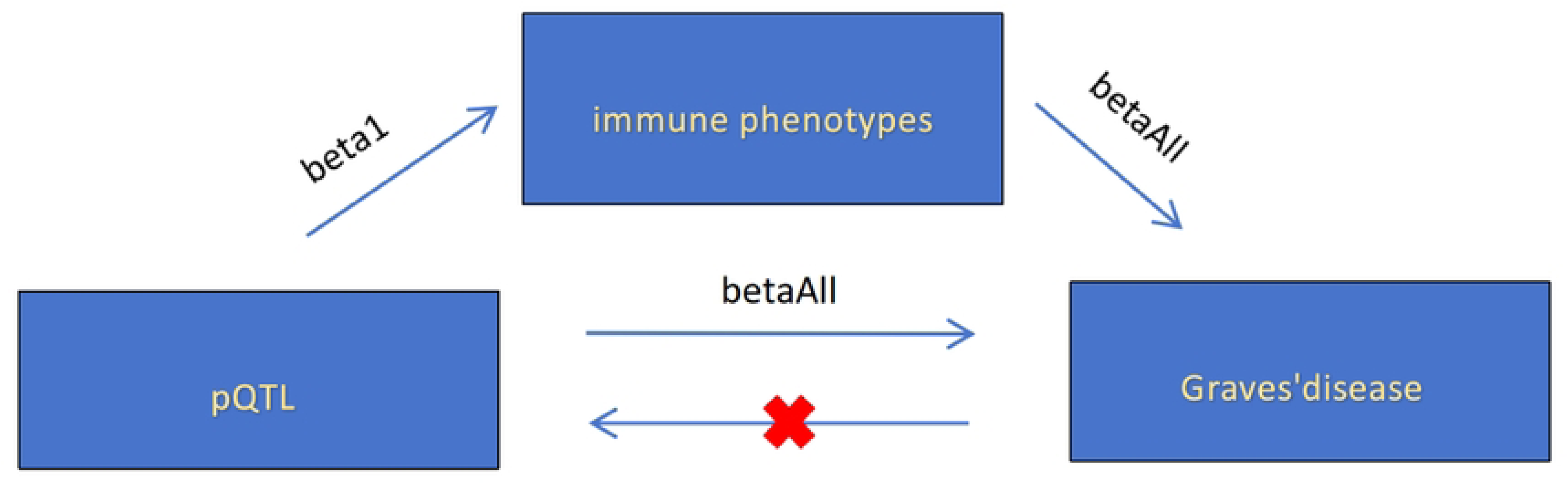
Sequential steps of the mediation analysis for pQTLs, immunophenotypes, and GD.

Additionally, based on the effect directions obtained through the pQTL-GD pathway (bataAll), the pQTL-immunophenotype pathway (beta1), and the immunophenotype-GD pathway (beta2), only mediation effects with logically consistent directions were retained. Specifically, if betaAll is positive, then beta1 and beta2 should have the same sign (both positive or both negative). Conversely, if the betaAll is negative, beta1 and beta2 should have opposite signs (one positive and the other negative). This logic ensures the mechanistic plausibility and directional consistency of the mediation effect.

The mediation effect, representing the indirect influence of pQTLs on GD through immunophenotypes, was derived by multiplying beta1 and beta2. Finally, the proportion of the mediated effect was calculated using the formula (beta1 × beta2) / betaAll[14].

To determine whether the identified proteins are potential targets of existing drugs or druggable genes, we analyzed their interactions with drugs using the DGIdb database. DGIdb integrates resources such as DrugBank, PharmGKB, and ChEMBL, supporting the search and filtering of drug-gene interactions and pharmacogenomic data.[15]

### The Impact of pQTL on GD

In the forward MR analysis, 36 results were identified with IVW p-values less than 0.05 using the Inverse Variance Weighted (IVW) method. Details are provided in S4 Table. No horizontal pleiotropy analysis was found due to an insufficient number of SNPs (fewer than 3). Consequently, 12 results were excluded because the number of SNPs was inadequate to ensure the robustness of the findings, as detailed in S5 Table. No heterogeneity was detected, as shown in S6 Table. The final results retained 24 pQTLs associated with GD. Notably, 10 pQTLs were identified as protective factors for GD (OR < 1), including FCRL1, POSTN, PMM2, GSTM3, APOF, HAPLN1, MTHFS, RNASET2, CST4, and APOL3. On the other hand, 14 pQTLs were identified as risk factors for GD (OR > 1), including F7, TIMD4, LRP11, AOC1, FCRL4, HAVCR1, GZMA, VNN2, ACE, DSG2, ENTPD5, VW2, DPEP1, and POFUT1, as detailed in Fig 2. Supporting visualizations, including forest plots, leave-one-out tests, and scatter plots, are provided in Files S1-S3.

### The Impact of Immune Phenotypes on GD

In the MR analysis between immune phenotypes and GD, 36 results had IVW p-values less than 0.05, as detailed in S7 Table. However, horizontal pleiotropy analysis revealed that HLA DR on CD14⁺ CD16⁻ monocytes and HLA DR on CD14⁺ monocytes had p-values less than 0.05, as shown in S8 Table, leading to the exclusion of these two results. In addition, the heterogeneity test identified 13 results with heterogeneity, as detailed in S9 Table. Using Weighted Median analysis, three results (IgD⁻ CD27⁻ AC, Naive DN (CD4⁻CD8⁻) %T cell, HLA DR on myeloid DC) were found to have p-values still less than 0.05, with effect directions consistent with those obtained through the IVW method. The remaining 10 results were excluded entirely. In the final set of 24 immune phenotypes, 12 were identified as protective factors for GD, and another 12 were considered to be associated with increased GD risk, as shown in Fig 3. Supporting visualizations, including forest plots, leave-one-out tests, and scatter plots, are provided in Files S4-S6.

### MR Analysis of Immune Phenotypes and pQTLs Reveals Mediation Effects

In the MR analysis targeting 24 immune phenotypes and their associations with pQTLs, we found that 19 pQTLs were associated with 11 immune phenotypes, as detailed in S10 Table. No horizontal pleiotropy or heterogeneity was detected, as shown in S11 Table and S12 Table. Reverse MR analysis on these 19 pQTLs did not reveal any evidence of reverse causation. After removing results with inconsistent effect directions, we ultimately identified that 8 pQTLs mediated 9 immune phenotypes, with the results summarized in Table 1.

Specifically, our analysis revealed that several immune cell phenotypes served as mediators in the relationships between specific pQTLs and GD. IgD⁻ CD27⁻ AC mediated the relationships of ACE, DPEP1, and PMM2 with GD with mediation proportions of 13.96%, 16.50%, and 7.98%, respectively. CD14 on CD33bright HLA-DR⁺ CD14dim mediated the relationship between PMM2 and GD with a mediation proportion of 11.67%. Naive DN (CD4⁻CD8⁻) %T cell mediated the relationship between FCRL4 and GD with a mediation proportion of 12.95%, while CD20 on IgD⁺ CD38⁻ unswitched memory B cells and CD45 on CD8bright T cells mediated 9.58% and 1.99%, respectively, of the relationship between FCRL4 and GD. CD39⁺ CD4⁺ %CD4⁺ mediated the relationships between POSTN and GD and ACE and GD with mediation proportions of 3.15% and 5.75%, respectively. CX3CR1 on CD14⁻ CD16⁻ mediated 2.74% of the relationship between CST4 and GD. Basophil %CD33dim HLA-DR⁻ CD66b⁻ mediated 7.54% of the relationship between HAPLN1 and GD. FSC-A on B cells mediated 6.98% of the relationship between FCRL1 and GD.

### Potential Drug Associations

According to DGIdb query results, potential drug associations were identified for five key pQTLs. Specifically, ACE is associated with several approved ACE inhibitors, including captopril, lisinopril, and enalapril. DPEP1 is linked to the approved drug cilastatin, as well as sorafenib and imatinib. CST4 and POSTN are associated with unapproved anti-serum (Antiserum), while PMM2 is linked to the unapproved drug EBSELEN. Detailed drug associations are provided in Table S13.

## Discussion

Our study indicates that ACE is positively correlated with GD by affecting the absolute number of IgD⁻CD27⁻ B cells.IgD⁻CD27⁻ B cells represent a distinct B cell subset. One study found that the proportion of IgD ⁻ CD27 ⁻ B cells was significantly increased in GD patients and was closely associated with thyroid-stimulating antibodies (TSAb) and TRAb. After treatment with antithyroid drugs, the proportion of these cells significantly decreased, correlating with improvements in thyroid function. This suggests that IgD ⁻ CD27 ⁻ B cells may serve as potential markers for treatment response, which is consistent with our findings [16].

Angiotensin-converting enzyme (ACE) is a key regulator of blood pressure and is known to be elevated in the peripheral blood of GD patients. However, previous studies have predominantly focused on its relationship with thyroid hormones [17–19]. Our research further highlights the significant role of ACE in the immune system, providing new insights into its functional implications. Studies have shown that in ACE-overexpressing mice, myeloid-derived suppressor cells (MDSCs) are reduced, while M1 macrophages are increased, enhancing the antigen-presenting ability of macrophages and promoting the activation of CD8+ and CD4+ T cells. The activation of CD4+ T cells further regulates B cells, leading to increased antibody production [20]. Our study similarly identifies the regulatory effects of ACE on both B and T cells (specifically CD39 on CD4+ T cells), which influences the development of GD.

The upregulation of CD39 on CD4+ T cells has been reported in various cancers, where its strong immunosuppressive function increases tumor risk [21–22]. This mechanism suggests that CD39 on CD4+ T cells may play a protective role in GD.

In our study, we discovered that CD14 on CD33bright HLA-DR+ CD14dim cells is positively correlated with GD and mediates the effect of PMM2 on GD. Furthermore, it is worth noting that we also observed a negative correlation between the proportion of CD33bright HLA-DR+ CD14-cells and GD.CD14, a core receptor for toll-like receptors (TLRs) [23], is highly expressed in the thyroid tissues of GD patients and is associated with an increased proportion of M1 macrophages, T follicular helper (Tfh) cells, and Th2 cells, potentially exacerbating GD [24,25].

The CD33bright HLA-DR+ cells are likely to be a subset of MDSCs [26]. MDSCs are a heterogeneous group of immune-suppressive cells that can suppress immune responses both under normal and pathological conditions [26]. However, the role of MDSCs in autoimmune diseases is complex. For instance, in a lupus mouse model, MDSCs inhibit CD4+ T cell proliferation through ARG1 and promote the expansion of regulatory B cells, exerting a protective effect [27]. On the other hand, studies have also shown that the frequency of MDSCs correlates with ARG1 activity, Th17 responses, and disease severity in systemic lupus erythematosus (SLE) patients [28]. Similar contradictory findings have been observed in rheumatoid arthritis (RA) [29, 27]. Our study further reveals that the role of CD33bright HLA-DR+ MDSC-like cells in GD may depend on the level of CD14 expression, potentially explaining the contradictory effects of MDSCs in autoimmune diseases.

This study is the first to reveal a positive correlation between PMM2 and GD. PMM2 is an enzyme involved in N-glycan synthesis and is widely expressed in immune cells. Deficiency of PMM2 can lead to reduced glycosylation of proteins, thereby affecting their normal functions, including the impairment of immune cell activity [30]. In patients with PMM2 activity deficiency, reduced neutrophil chemotaxis and weakened humoral immune responses following vaccination have been observed [31]. Additionally, abnormal glycosylation of B cells may impact the activity of proteins such as CD22, thereby modulating B cell function [32]. Studies have shown that IgG N-glycosylation patterns in GD patients undergo dynamic changes before and after immunosuppressive treatment. These changes not only serve as potential biomarkers for assessing the disease state but may also be closely associated with treatment outcomes [33]. Furthermore, PMM2 deficiency in patients leads to abnormal glycosylation of GPI-anchored proteins like CD16 and CD14 in neutrophils and monocytes [34]. Thus, based on our findings, we hypothesize that PMM2 influences immune cells such as B cells (e.g., IgD⁻ CD27⁻ B cells) and certain immune cells (e.g., CD14 in CD33bright HLA-DR+ CD14dim cells) through glycosylation, subsequently impacting the pathogenesis of GD.

FCRL4 is a key regulator of B cell function, inhibiting BCR signaling to suppress B cell proliferation while enhancing TLR signaling to promote inflammation [35]. Previous studies have shown that FCRL4 is highly expressed in the peripheral blood B cells of GD patients and is associated with hyperthyroidism, but it is not correlated with TSH receptor antibody (TSHRAb) positivity, and its expression does not significantly change in GO patients [36, 37]. It was previously thought that FCRL4 does not directly regulate TSHRAb production in GD, and the specific mechanisms remain unclear. Our study further suggests that FCRL4 may influence GD by modulating the immune response of both B and T cells.

FCRL4 is highly expressed in memory B cells, including IgD⁺CD38⁻unswitched memory B cells [38], which belong to a specific subset of memory B cells. CD20 is known to enhance B cell activation, thereby amplifying immune responses [39]. Compared with FCRL4 ⁻ memory B cells, CD20 is highly expressed in FCRL4+ memory B cells, which do not express CD20 in plasma cells [38]. Moreover, FCRL4+ memory B cells are less likely to differentiate into plasma cells [40]. We propose that FCRL4 may prevent the premature differentiation of unswitched memory B cells into plasma cells by negatively regulating BCR signaling, thereby maintaining the number of IgD⁺ CD38 ⁻ cells and enhancing their pro-inflammatory function through CD20 expression. Additionally, the enhanced TLR9 signaling in these B cells may further augment their inflammatory potential.

Furthermore, we identified that FCRL4 regulates certain T cell subsets. Naive DN (CD4⁻CD8⁻) T cells, which secrete pro-inflammatory cytokines such as IL-17 and IFN-γ, play critical roles in various autoimmune diseases and have been observed in both mouse models and human studies [41, 42].

CD45 is a protein tyrosine phosphatase broadly expressed on the surface of immune cells, regulating the phosphorylation of LCK and dynamically controlling the strength of TCR signaling in CD8+ T cells. CD8+ T cells with high CD45 expression exhibit weaker TCR responses to self-antigens, thereby preventing excessive immune reactions [43]. The interaction between FCRL4 and these T cells may play a crucial role in immune regulation in GD, although the precise mechanisms require further investigation.

DPEP1 is an enzyme that facilitates immune responses by promoting the recruitment and adhesion of neutrophils in inflammatory environments. In an LPS-induced inflammation model, DPEP1 deficiency was found to reduce the adhesion of neutrophils in the lungs and liver [44].

Additionally, in a model of renal ischemia-reperfusion injury (IRI), DPEP1 antagonists prevented the recruitment of neutrophils and monocytes, thereby alleviating inflammation and acute kidney injury (AKI) [45]. In our study, we observed that DPEP1 positively regulates B cells (IgD ⁻ CD27⁻ AC), thereby contributing to the development of GD.

In the following analysis, based on the identified protein-drug interactions, we further screened potential drug candidates for GD.

Studies have shown that ACE inhibitors (ACEIs) exhibit significant anti-inflammatory effects in various models of inflammation and autoimmune diseases. For example, Lisinopril has been found to alleviate symptoms of experimental thyroid granulomatous thyroiditis by reducing the production of TGF-β1 and decreasing pro-inflammatory cytokines and collagen deposition [46]. Captopril can reduce IFN-γ production and activate suppressor of cytokine signaling (SOCS)-1 and SOCS-3, thereby attenuating inflammation in the experimental autoimmune encephalomyelitis (EAE) model [47]. Additionally, in lupus nephritis models, ACEIs effectively protect the kidneys by lowering Th2-associated cytokines TGF-β 1 and TGF-β 2 levels [48]. Furthermore, ACEIs can inhibit NF-κ B activation, preventing vascular inflammation associated with atherosclerosis [49]. These studies collectively highlight the potential of ACEIs in modulating immunity and exerting anti-inflammatory effects in various immune-related diseases.

Based on the integrated analysis of our study, we suggest further exploration of the role of ACE inhibitors in the immune modulation and inflammation control in GD. Assessing their potential efficacy could lead to novel therapeutic strategies for this disease.

Cilastatin, an FDA-approved DPEP1 inhibitor, presents another promising candidate for future research. Further investigations are needed to explore its potential mechanisms in GD, particularly whether modulation of DPEP1 activity can improve metabolic dysregulation or pathological features associated with GD. Such studies would provide crucial evidence for evaluating DPEP1 as a key therapeutic target for GD, and help assess the clinical application prospects of cilastatin and similar inhibitors in the treatment of GD.

This study is the first to combine pQTLs with immune cell-mediated mechanisms to explore their impact on the onset and progression of GD. Additionally, it represents the first Mendelian randomization (MR) study that integrates pQTLs and immune cell mechanisms, making it both innovative and pioneering. Through this comprehensive approach, we not only validated previously identified GD-related pQTLs but also discovered novel pQTLs and potential targets, thus expanding our understanding of the pathogenesis of GD and other autoimmune diseases.

The findings of this study have significant scientific and clinical implications. On the one hand, they reveal several potential pathogenic mechanisms, providing new insights into the complex pathology of GD. On the other hand, by combining pQTLs with immune cell targets, our research strategy not only opens new avenues for drug development but also lays the theoretical foundation for combination targeted therapies. In the future, drug development targeting these pathways, either individually or in combination, could significantly improve the precision and effectiveness of treatment for GD and related autoimmune diseases.

However, while acknowledging the strengths of this study, its limitations must also be considered. As an exploratory study, this research aimed to identify potential causal pathways and candidate signals, providing a foundation for subsequent validation studies. Therefore, corrections for multiple comparisons were not performed in order to maximize the discovery of candidate signals. However, this approach may increase the risk of false positives, which will require rigorous validation in future studies to ensure the reliability of the results. Additionally, heterogeneity was observed in the MR analysis of immune phenotypes and GD, which may be attributed to population differences between Icelandic and European cohorts. Although a weighted median method was applied to address the heterogeneity, caution is still required in interpreting the results.

## Data availability

The original contributions presented in the study are included in the article.

## Acknowledgments

We sincerely thank the deCODE Genetics study, FinnGen, and the GWAS Catalog for providing the data resources that were integral to this study

## Supporting information

**S1 File Forest plot for MR results between pQTLs and Graves’ disease**

**S2 File Leave-one-out plot for MR results between pQTLs and Graves’ disease**

**S3 File Scatter plot for MR results between pQTLs and Graves’ disease**

**S4 File Forest plot for MR results between immunophenotypes and Graves’ disease**

**S5 File Leave-one-out plot for MR results between immunophenotypes and Graves’disease**

**S6 File Scatter plot for MR results between immunophenotypes and Graves’ disease**

**S7 File Forest plot for MR results between pQTLs and immunophenotypes**

**S8 File Leave-one-out plot for MR results between pQTLs and immunophenotypes.**

**S9 File Scatter plot for MR results between pQTLs and immunophenotypes**

**S1 Table The instrumental variables for pQTL.**

**S2 Table The instrumental variables for immunophenotypes**

**S3 Table The instrumental variables for Graves’ disease**

**S4 Table The ivw results with p-values less than 0.05 between pQTLs and GD**

**S5 Table The horizontal pleiotropy between pQTLs and Graves’ disease**

**S6 Table The heterogeneity between pQTLs and Graves’ disease**

**S7 Table The ivw results between immunophenotypes and Graves’ disease**

**S8 Table The horizontal pleiotropy between immunophenotypes and Graves’ disease**

**S9 Table The heterogeneity between immunophenotypes and Graves’ disease**

**S10 Table The ivw results between pQTLs and immunophenotypes**

**S11 Table The horizontal pleiotropy between pQTLs and immunophenotypes**

**S12 The heterogeneity between pQTLs and immunophenotypes**

**S13 Table Results of the related drug query**

**S14 Table STROBE-MR guidance**

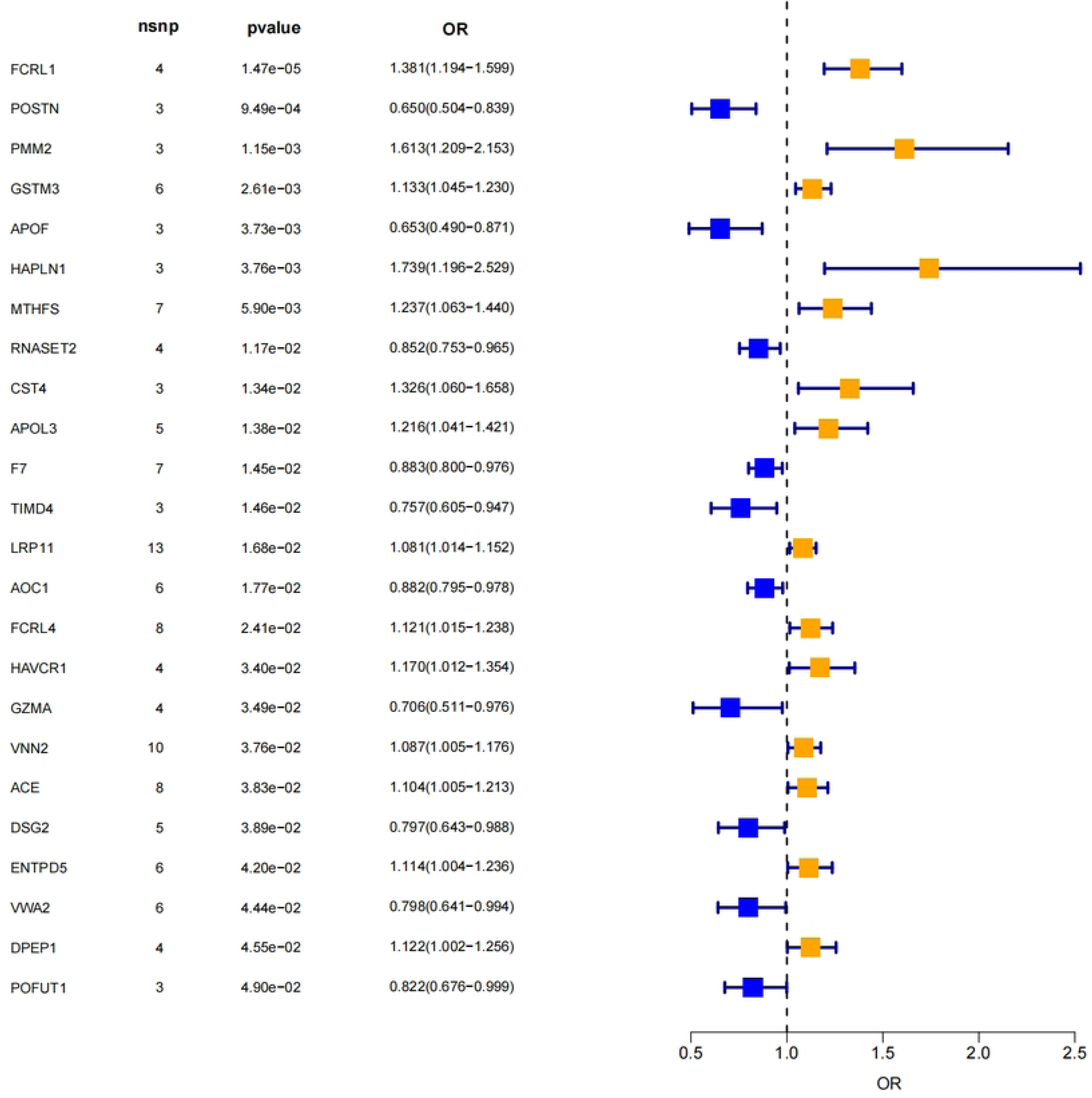

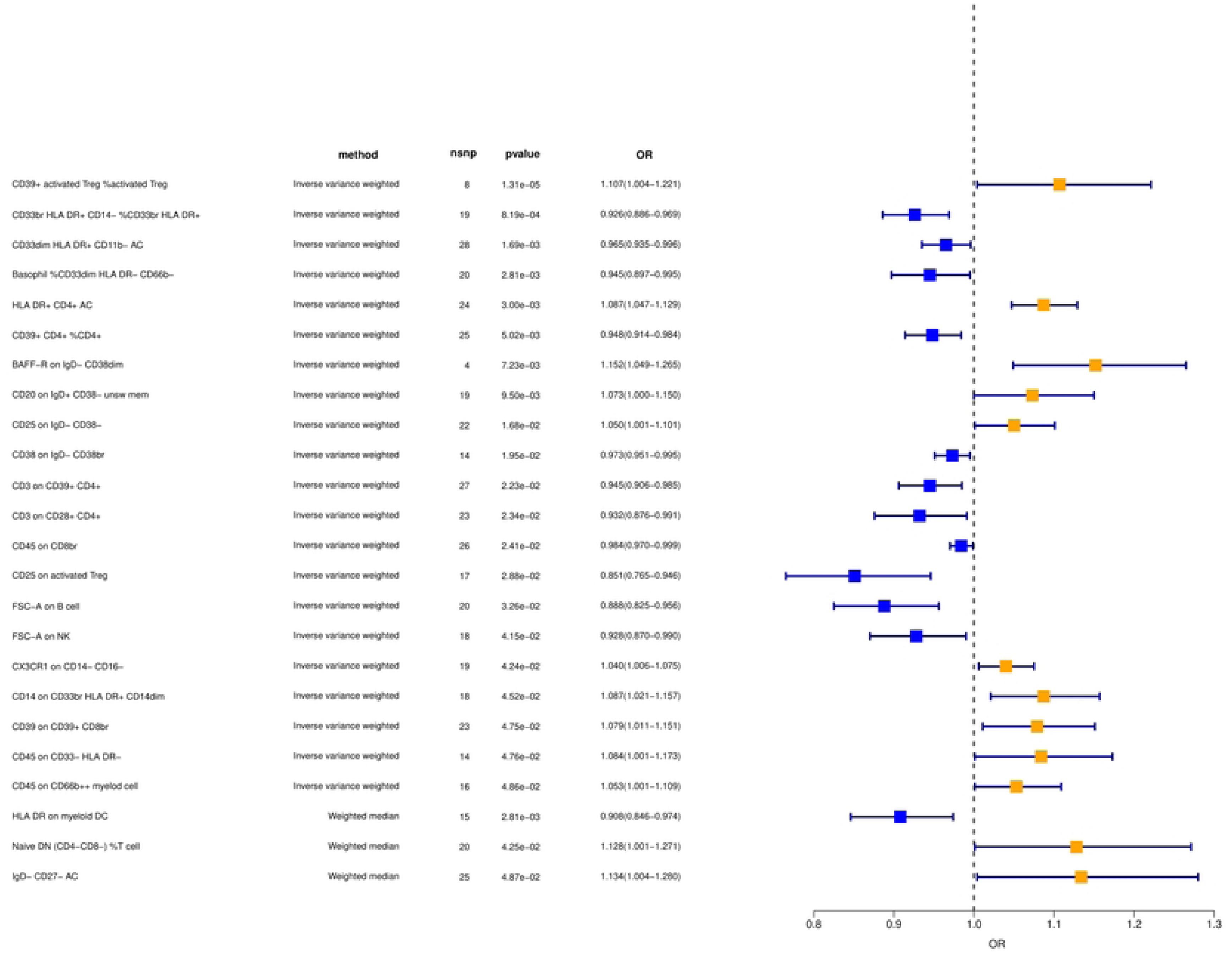

